# Comparison of multiple whole-genome and *Spike*-only sequencing protocols for estimating variant frequencies via wastewater-based epidemiology

**DOI:** 10.1101/2022.12.22.22283855

**Authors:** Lucy A. Winder, Paul Parsons, Gavin Horsburgh, Kathryn Maher, Helen Hipperson, Claudia Wierzbicki, Aaron R. Jeffries, Mathew R. Brown, Aine Fairbrother-Browne, Hubert Denise, Mohammad S. Khalifa, Irene Bassano, Ronny van Aerle, Rachel Williams, Kata Farcas, Steve Paterson, Paul G. Blackwell, Terry Burke

## Abstract

Sequencing of SARS-CoV-2 in wastewater provides a key opportunity to monitor the prevalence of variants spatiotemporally, potentially facilitating their detection simultaneously with, or even prior to, observation through clinical testing. However, there are multiple sequencing methodologies available. This study aimed to evaluate the performance of alternative protocols for detecting SARS-CoV-2 variants. We tested the detection of two synthetic RNA SARS-CoV-2 genomes in a wide range of ratios and at two concentrations representative of those found in wastewater using whole-genome and *Spike*-gene-only protocols utilising Illumina and Oxford Nanopore platforms. We developed a Bayesian hierarchical model to determine the predicted frequencies of variants and the error surrounding our predictions. We found that most of the sequencing protocols detected polymorphic nucleotide frequencies at a level that would allow accurate determination of the variants present at higher concentrations. Most methodologies, including the *Spike*-only approach, could also predict variant frequencies with a degree of accuracy in low-concentration samples but, as expected, with higher error around the estimates. All methods were additionally confirmed to detect the same prevalent variants in a set of wastewater samples. Our results provide the first quantitative statistical comparison of a range of alternative methods that can be used successfully in the surveillance of SARS-CoV-2 variant frequencies from wastewater.

**Impact:** Genetic sequencing of SARS-CoV-2 in wastewater provides an ideal system for monitoring variant frequencies in the general population. The advantages over clinical data are that it is more cost efficient and has the potential to identify new variants before clinical testing. However, to date, there has been no direct comparison to determine which sequencing methodologies perform best at identifying the presence and prevalence of variants. Our study compares seven sequencing methods to determine which performs best. We also develop a Bayesian statistical methodology to estimate the confidence around variant frequency estimates. Our results will help monitor SARS-CoV-2 variants in wastewater, and the methodology could be adapted for other disease monitoring, including future pandemics.

## Introduction

The regular testing of wastewater for the presence of SARS-CoV-2 using quantitative PCR (qPCR) has been widely adopted in response to the Covid-19 pandemic since 2020 in the UK, including at major sewage treatment works (Farkas et al., 2020; Larsen and Wigginton, 2020). This monitoring tool has proven to be a valuable adjunct to other data sources on the progress, prevalence and location of Covid-19 in the human population within the catchment of each sampling site. Wastewater monitoring, therefore, provides both temporal and spatial information on the development of the Covid-19 pandemic. Furthermore, this methodology is unbiased by asymptomatic infections (Sah et al. 2021) and could potentially allow for the detection of variants before clinical cases have presented (Peccia et al 2020). Early detection of new variants in the UK has numerous public health benefits, giving policymakers and healthcare professionals more time to prepare for new Covid-19 infection waves.

The power of qPCR is accurately determining the presence, absence and concentration of SARS-CoV-2, and, on occasion, via protocol modification, alternative focal variants (Alcoba-Florez et al 2020; Kudo et al 2020). However, qPCR assays that only target one genomic region can be susceptible to false negatives when the sample is either high in inhibitors, degraded and/or of a concentration outside the assays limit of detection (Forootan et al. 2017; Schrader et al. 2012; Bahreini et al. 2020). Variants can also lead to false negatives in qPCR assays, if a mutation has emerged at the primer binding site (Lefever et al. 2013). This issue was highlighted with the emergence of the B.1.1.7 variant; the deletion H69-V70 falls in the target region of a primer set run routinely, leading to the complete dropout of the S marker from tests (Bal et al. 2021; Volz et al. 2021). The most powerful method to detect variants is potentially through RNA sequencing.

Next-generation sequencing (NGS) technologies, especially those with short read lengths, have worked effectively even on degraded samples (Sanz and Köchling 2019; Burrell et al. 2015). This is because the short read length increases the chance of successfully generating amplicons in fragmented samples (Burrell et al. 2015; Berglund et al. 2011) and the large number of reads means that even rare sequences within a sample can be identified (Ryu et al. 2018). The RNA of SARS-CoV-2 can be detected in the faeces of infected human hosts (Chen et al. 2020) and can persist in aquatic environments for several days (Bivins et al. 2020; Sala-Comorera et al. 2021). Furthermore, SARS-CoV-2 sequencing data from wastewater samples can be used to determine variants and their frequency, with evidence suggesting this is more sensitive than clinical surveillance (Karthikeyan et al. 2022; Morvan et al. 2022). However, there has been, to date, no formal comparison of alternative NGS protocols in detecting SARS-CoV-2 in wastewater. Determining the methods which are better able to determine variant frequencies, particularly at low concentrations, could be invaluable to efforts monitoring SARS-CoV-2 in wastewater.

This study aimed to compare alternative sequencing protocols to identify the most efficient for sequencing mixtures of SARS-CoV-2 variants and estimate the frequencies of those present. The protocols were tested initially by creating mixtures of synthetic RNA for two variants that reached high frequencies in the course of the pandemic in the UK. Two concentrations of synthetic RNA were designed to be comparable to those seen in wastewater. The study included the development of a Bayesian statistical approach to attach credible intervals to variant frequency estimates. The analysis also included a comparison between qPCR analysis of specific variants and PCR-based SNP detection of variants. Finally, the methods were compared through the sequencing of RNA obtained from wastewater collected from a population experiencing a high level of Covid-19 infection.

## Methods

### Mixtures of synthetic SARS-CoV-2 RNA

We obtained two synthetic SARS-CoV-2 RNA genomes from Twist Bioscience (South San Francisco, CA): Control C12 (B.1.369; GenBank EPI_ISL_420244; GISAID England/SHEF-C05B2/2020; denoted “SHEF”) and Control 15 alpha (B.1.1.7; EPI_ISL_601443; England/MILK-9E05B3/2020; denoted “MILK”). These were each supplied as six contiguous *ca* 5-kb fragments at an approximate concentration of 10^6^ genome copies (gc) per microlitre. Any amplicons designed across a breakpoint between adjacent RNA sequences could not be amplified. We prepared a range of mixtures at nominal concentrations of 200 gc/µL and 20 gc/µL in a range of ratios from 100% SHEF to 100% MILK (Table 1). These two concentrations were chosen to be comparable to the amounts of SARS-CoV-2 RNA obtained from 250-mL wastewater samples in the UK national monitoring programme (corresponding to concentrations of 10^2^–10^5^ gc/l, UKHSA 2021). We note that the number of genome copies of a minor variant is expected to be limiting when present at a low proportion in a sample of low overall concentration; for example, a variant at 1% frequency in the lower concentration used here is only expected to be present, on average, as a single copy in a 5-µL PCR reaction (as used by several of the methods tested here).

**Table 1:** Estimated concentrations of synthetic RNA variants in mixtures, corrected using sequencing data (see text).

| Mixture no. | Nominal proportions | | Corrected proportions | | High concentration/<br>Estimated genome copies per μl | | Low concentration/<br>Estimated genome copies per μl | |
| --- | --- | --- | --- | --- | --- | --- | --- | --- |
|  | SHEF | MILK | SHEF | MILK | SHEF | MILK | SHEF | MILK |
| 1 | 0.000 | 1.000 | 0.000 | 1.000 | 0.0 | 158.4 | 0.0 | 15.8 |
| 2 | 0.010 | 0.990 | 0.015 | 0.985 | 2.4 | 156.8 | 0.2 | 15.7 |
| 3 | 0.050 | 0.950 | 0.074 | 0.926 | 12.1 | 150.5 | 1.2 | 15.0 |
| 4 | 0.100 | 0.900 | 0.145 | 0.855 | 24.2 | 142.6 | 2.4 | 14.3 |
| 5 | 0.200 | 0.800 | 0.276 | 0.724 | 48.3 | 126.7 | 4.8 | 12.7 |
| 6 | 0.500 | 0.500 | 0.604 | 0.396 | 120.8 | 79.2 | 12.1 | 7.9 |
| 7 | 0.800 | 0.200 | 0.859 | 0.141 | 193.3 | 31.7 | 19.3 | 3.2 |
| 8 | 0.900 | 0.100 | 0.932 | 0.068 | 217.4 | 15.8 | 21.7 | 1.6 |
| 9 | 0.950 | 0.050 | 0.967 | 0.033 | 229.5 | 7.9 | 23.0 | 0.8 |
| 10 | 0.990 | 0.010 | 0.993 | 0.007 | 239.2 | 1.6 | 23.9 | 0.2 |
| 11 | 1.000 | 0.000 | 1.000 | 0.000 | 241.6 | 0.0 | 24.2 | 0.0 |
| 12 | 0.000 | 0.000 | 0.000 | 0.000 | 0 | 0 | 0 | 0 |

The SNP frequencies obtained from the sequencing data indicated that the two concentrated RNAs differed in initial concentration. We therefore used the numbers of sequencing reads obtained for diagnostic SNPs in the *Spike* gene for the nominal 50:50 variant mixes to correct the ratios (using data for two replicate sequencing runs obtained using each of the Oxford Nanopore and Illumina sequencing SubARTIC methods, described below, and assuming that the mean starting concentration of the two variants was 10^6^ gc/ul), and from these data we calculated corrected estimated ratios in the utilised mixtures (Table 1).

### Wastewater Sample Collection, Concentration and Extraction

Wastewater grab samples (1 L per sample) were collected from 17 locations across the London sewer network on five consecutive days from the 10–14 January 2021 as part of the ongoing Environmental Monitoring for Health Protection (EMHP) programme in England. Samples were transported to Eurofins BioPharma, Glastrup, Denmark and stored at 4–6°C until RNA extraction, minimising RNA degradation. Two hundred-millilitre subsamples from each location were pooled on each day (totalling 3.4 L), mixed, then split into 20 × 100-mL subsamples and then purified via centrifugation (10,000 X*g* for 30 minutes at 4°C). Fifty millilitres of each supernatant was retained, with the pH adjusted (to 7.0–7.6 using 1 M NaOH) prior to concentration into 2 mL using polyethylene glycol precipitation (PEG, 40% PEG 8000,118% NaCl) overnight at 4°C followed by further centrifugation (10,000 X*g* for 30 minutes at 4°C). RNA was then extracted using the VIRSeek RNAExtractor kit (Eurofins Technologies, Germany) and the KingFisher Flex Purification System (Thermo, UK)⍰1according to the manufacturers’ instructions, so generating 20 100-µL RNA extracts per date. For each date, the RNA extracts were pooled, mixed and re-aliquoted into 20 × 100-µL extracts, which were stored at -20°C until distribution to the participating laboratories for sequencing.

### cDNA Synthesis and Sequencing

#### (1) ARTIC Illumina

The ARTIC protocol has been widely implemented for sequencing clinical samples of SARS-CoV-2 on the Oxford Nanopore platform (Quick 2020; examples of use: Rivett et al 2020; Tegally et al 2021). This protocol uses a primer set (version 3 here; Quick 2020) that produces amplicons across the whole genome (Table 2), and was adapted in Liverpool so that the amplicons could be sequenced on the Illumina MiSeq platform. The primers are tiled, with even and odd amplicons amplified separately before pooling for sequencing library preparation. Here, we used the protocol to sequence mixtures of synthetic SARS-CoV-2 RNA and RNA recovered from wastewater on the Illumina MiSeq instrument.

**Table 2:**
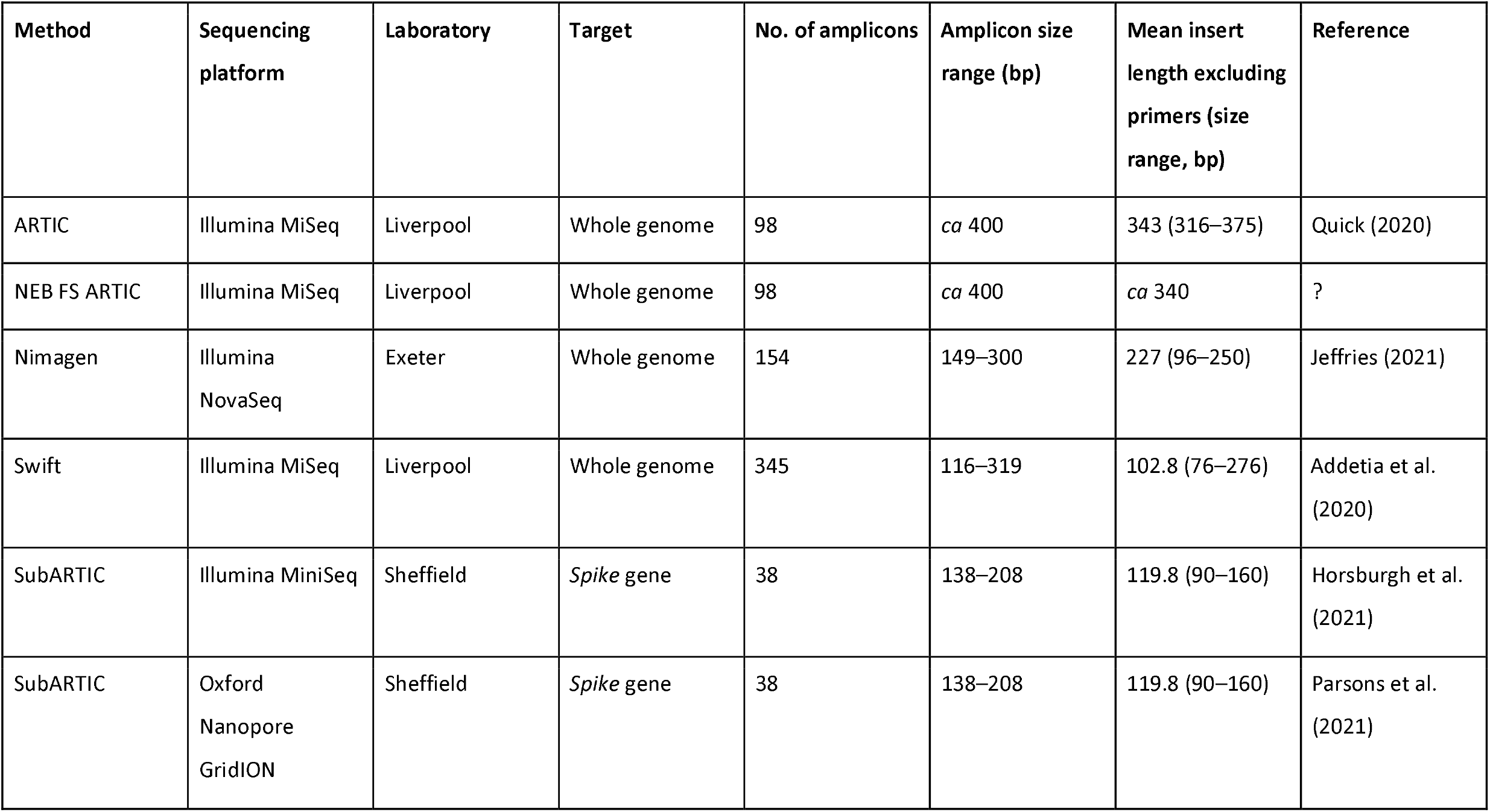
Sequencing / genotyping methods assessed for their capacity to estimate variant frequencies.

| Method | Sequencing platform | Laboratory | Target | No. of amplicons | Amplicon size range (bp) | Mean insert length excluding primers (size range, bp) | Reference |
| --- | --- | --- | --- | --- | --- | --- | --- |
| ARTIC | Illumina MiSeq | Liverpool | Whole genome | 98 | *ca* 400 | 343 (316–375) | Quick (2020) |
| NEB FS ARTIC | Illumina MiSeq | Liverpool | Whole genome | 98 | *ca* 400 | *ca* 340 | ? |
| Nimagen | Illumina NovaSeq | Exeter | Whole genome | 154 | 149–300 | 227 (96–250) | Jeffries (2021) |
| Swift | Illumina MiSeq | Liverpool | Whole genome | 345 | 116–319 | 102.8 (76–276) | Addetia et al. (2020) |
| SubARTIC | Illumina MiniSeq | Sheffield | *Spike* gene | 38 | 138–208 | 119.8 (90–160) | Horsburgh et al. (2021) |
| SubARTIC | Oxford Nanopore GridION | Sheffield | *Spike* gene | 38 | 138–208 | 119.8 (90–160) | Parsons et al. (2021) |

RNA extracted from wastewater was DNase-treated to remove residual DNA to prevent PCR inhibition; 40 ul RNA was treated using the TURBO DNA-free Kit (Ambion). Following DNase treatment, the RNA was purified and concentrated with a 1.8x RNA bead clean up and eluted in 16 µl nuclease-free water. Twist synthetic RNA standards were used directly. Reverse transcription was performed in duplicate using 2 µl NEB Lunascript and 8 µl RNA, including negative and positive controls, with the cycling conditions as follows: 25°C for 2 minutes, 55°C for 10 minutes, 95°C for 1 minute. Tiling PCR using the ARTIC v3 primer sets was performed using 4 µl cDNA input per PCR. The cycling conditions were initial denaturation at 98°C for 30 seconds, followed by 30 cycles of denaturation at 98°C for 15 seconds, and annealing and extension at 63°C for 5 minutes, with a final hold at 4°C. Following PCR, amplicon pools A and B for each sample were pooled and a 1:1 Ampure XP bead (Beckman) purification was performed, and eluted in 20 µl nuclease-free water. Ten microlitres of purified library was used for library preparation.

The library was prepared for sequencing using a one-third volume NEB Next Ultra II protocol and indexed using unique dual indexes (IDT), using the following cycling conditions: initial denaturation at 98°C followed by 5 cycles of denaturation at 98°C for 30 seconds, and annealing and extension at 65°C for 75 seconds, a final extension at 65°C for 5 minutes and a final hold at 4°C. The indexed library was pooled without normalisation, taking 2 µl of each sample, and purified using a 0.8x Ampure purification. The final library was quantified and run on the Agilent Bioanalyzer. The library concentration was determined using qPCR prior to sequencing using the Illumina Miseq v2 250 × 250 cycle kit.

#### (2) NEB FS ARTIC Illumina

RNA extracted from wastewater was DNase-treated to remove residual DNA in order to prevent PCR inhibition; 40 µlRNA was treated with the TURBO DNA-free Kit (Ambion). Following DNase treatment, the RNA was purified and concentrated with a 1.8x RNA bead clean up and eluted in 16 µlnuclease-free water. Twist synthetic RNA standards were used directly. Reverse transcription was performed in duplicate using 2 µlNEB Lunascript and 8 µlRNA, including negative and positive controls, with the cycling conditions as follows: 25°C for 2 minutes, 55°C for 10 minutes, 95°C for 1 minute. Tiling PCR using the ARTIC v3 primer pools was performed using 4 µlcDNA input per reaction with the following cycling conditions: initial denaturation at 98°C for 30 seconds, followed by 30 cycles of denaturation at 98°C for 15 seconds and annealing and extension at 63°C for 5 minutes, with a final hold at 4°C. Following PCR, amplicon pools A and B for each sample were pooled and a 1:1 Beckman Ampure XP bead purification was performed, and eluted in 20 µlnuclease-free water.

Ten microlitres of pooled purified PCR product was fragmented using the NEBNext Ultra II FS DNA Library Prep Kit, following the protocol for inputs ≤100 ng. The library was fragmented for 30 minutes at 37°C to fragment amplicons to ∼120 bp. Adapter ligated libraries were purified using a 0.9x Ampure bead clean up, and eluted in 8 µlnuclease-free water and indexed using unique dual indexes (IDT) with the following cycling conditions: 98°C for 30 seconds, followed by 5 cycles of denaturation at 98°C for 30 seconds and annealing and extension at 65°C for 75 seconds, with a final extension at 65°C for 5 minutes. The indexed library was pooled without normalisation, taking 2 µlof each sample and purified using a 1:1 Ampure purification followed by a 0.8x purification to remove remaining short fragments. Libraries were run on the Agilent Bioanalyzer and quantified using qPCR prior to sequencing. Sequencing was performed using Illumina Miseq v2 150 × 150 cycle kit.

#### (3) NimaGen Illumina

The EasySeq SARS-CoV-2 WGS Library Prep Kit (NimaGen, Nijmegen, Netherlands) protocol has been implemented for large-scale sequencing of wastewater in the UK (Jeffries 2021). We used version 2 in this study.

Twenty microlitres of extracted RNA was cleaned up using 1.8 X Mag-Bind Total Pure NGS Cleanup beads and eluted in 9 µL of ultrapure water. Reverse transcription was then performed on 8 µL of eluted RNA using Lunascript (New England Biolabs, Ipswich, MA, USA) at 25°C for 2 minutes, 55°C for 45 minutes and 95°C for 1 minute, followed by a 4°C holding temperature. cDNA (2.5 µL) and 13.5 µL mastermix (New England Biolabs; i.e., mixture of reagents at concentrations optimal for PCR preparation) were then added to each of the two PCR plates from the Nimagen SARS-CoV-2 kit and plated on a PCR thermal cycler for the recommended cycling conditions. The following day, 3.5 µL of each reaction in a plate was pooled together into a 1.5-ml tube corresponding to each plate and a matching volume of T0.1E (10 mM Tris-HCl pH 8.0, 0.1 mM EDTA) was added. Nimagen’s Ampliclean beads at 0.85x were then added, mixed and left to sit at room temperature for 5 minutes. After two ethanol washes, the library was eluted in 100 µL of T0.1E. An additional 0.85x Ampliclean bead cleanup was performed and the purified library eluted in 25 µL of T0.1E. Final DNA concentrations were then determined by Qubit fluorometry and the readings entered into a molarity calculator provided by Nimagen. Pooled libraries containing Unique Dual Indexes (UDIs) were then loaded on the NovaSeq SP 300 flow cell in a 2 × 150-bp read format, spiked with 5% PhiX.

#### (4) Swift Illumina

Wastewater and synthetic samples were reverse transcribed using LunaScript with a 20-minute incubation time. Libraries were generated using the Swift Normalase Amplicon Panels (SNAP) SARS-CoV-2 Additional Genome Coverage, following the kit protocol. Amplicons were indexed using the SNAP Unique Dual Indexing Primer Plates. Optimal normalisation using Normalase was omitted due to low yields. Instead, libraries were run on the Agilent Fragment Analyzer and equimolar pooled. The final pooled library was quantified using Qubit and qPCR. Sequencing was performed using Illumina MiSeq v2 150 × 150 cycle kit.

#### (5) SubARTIC *Spike* Sequencing

We designed a sequencing protocol for the *Spike gene* region of SARS-CoV-2 by modifying the ARTIC protocol (above). The protocol used a redesigned primer set (version 3.2, Horsburgh et al. 2021, Parsons et al. 2021) where the amplicons had a reduced size range of 141–208 bp. The primers were tiled, with even and odd amplicons amplified separately before pooling for preparing sequencing libraries.

In brief, cDNA was synthesised from each RNA sample and a negative control (molecular grade water) using Lunascript (New England Biolabs, Ipswich, MA). This method does not require RNA purification. The primers are split into two tiled pools, even and odd, and PCR amplified in separate reactions. For each reaction, 4.5 µl cDNA was combined with 6.25 µl Q5 Hotstart High fidelity 2x Mastermix (New England Biolabs, Ipswich, MA) and 1.75 µl of the primer pool (10 mM). PCR products were then pooled and a second negative control sample (molecular grade water) included before sequencing the PCR amplicons on Day 2.

##### (a) Illumina

We used the SubARTIC sequencing v 3.2 protocol before loading the libraries onto an Illumina MiniSeq sequencer in Sheffield, as described in detail by Horsburgh et al. (2021). Up to sixty-six samples were included in a single sequencing run, including two negative controls. We added 35µlof PhiX at 1.4 pM to 500 µl of the 1.4 pM library before running on the MiniSeq. Sequencing produced 2x 150-bp paired-end reads.

##### (b) Oxford Nanopore

Much of the sequencing to identify SARS-CoV-2 in clinical samples has been undertaken using Oxford Nanopore Technology instruments (using the ARTIC protocol; see (1) above). SubARTIC v 3.2 sequencing of the synthetic mixes and wastewater samples was implemented here on the Oxford Nanopore GridION platform. A detailed protocol is provided by Parsons et al. (2021). Libraries were prepared using a modified Amplicon by Ligation protocol (SQK-LSK109; Oxford Nanopore Technologies, Oxford, UK) with Native Barcoding (EXP-NBD104, EXP-NBD114; Oxford Nanopore Technologies) and run for 18 hours on an R.9.4.1 flow cell using an Oxford Nanopore GridION sequencer. High accuracy basecalling was used with a minimum barcoding score of 80. All other parameters were set to their default values.

### Bioinformatics

Sequencing data were analysed using a modified version of the nCoV2019-ARTIC pipeline, which was originally developed for the analysis of clinical samples of SARS-CoV-2 (https://artic.network/ncov-2019/ncov2019-bioinformatics-sop.html). Briefly, reads were first mapped to the SARS-CoV-2 reference genome (NCBI Genbank Accession MN908947.3) with BWA V0.7.17 (Li, 2013). iVar (Grubaugh et al., 2019) was used to remove the primers based on positional information. An additional primer trimming step, using Cutadapt v1.18 (Martin, 2011), was included for the SubARTIC dataset. SNPs and Indels were identified using VarScan v2.3 (Koboldt et al., 2012) using a *p*-value threshold of 0.01. For simplicity, and comparability between Illumina and Oxford Nanopore instruments, given that the detection of indels is less reliable using the latter, we excluded indels in the analysis of the synthetic variant data.

### Statistical Analysis

In wastewater samples containing SARS-CoV-2 RNA, variants are often at very low concentration and will vary in proportion from 0–100%. In such RNA mixtures where one variant is present at a low proportion, detection of the associated diagnostic SNPs can be stochastic. We therefore developed a probabilistic model to quantify these errors and relate the data from all the SNPs associated with a variant to the frequency of that variant, and used Bayesian statistical methods to estimate its parameters based on the data from the synthetic RNA mixtures.

The model expresses the mean observed proportions of reads containing SNP-defining variants as a function of the true proportions of variants; this function has a cubic form, based on empirical observations of the synthetic data, with two parameters capturing the error level at low concentrations and the curvature of the function. Because errors can occur at various stages in the processing of the samples, there is dependence between reads, which means that the variability in counts greatly exceeds the binomial variation that would follow from independent reads. A standard approach would be to accommodate this dependence by allowing variation in the mean between SNPs, typically using a beta-binomial distribution. For the present data, a beta-binomial distribution did not allow sufficiently heavy-tailed distributions for the observed counts; instead, for each SNP we used a two-component mixture of beta-binomial distributions, each with the mean determined by the ‘cubic’ function. This entails a further three parameters, defining the variances of each component and their relative weighting. Reads for different SNPs are taken to be independent, given the parameters. Further details are given in the supplementary material (SI Appendix 1).

To estimate the parameters of the model for each sequencing method, we used a Bayesian statistical approach to fit the model to the synthetic samples containing known proportions of the two variants. The model-fitting used a Markov chain Monte Carlo algorithm as implemented in the JAGS package (Plummer, 2017, 2021a), run via the R package *rjags* (Plummer, 2021b). Uninformative prior distributions were used for the model parameters, separately for each method. Again, further details are given in the supplementary material (SI Appendix 1). The joint posterior distribution for the parameters in each case is complex and highly dependent, and so was represented for further analysis by a large Monte Carlo sample produced from JAGS, rather than attempting to summarise it parametrically.

To apply the model to data with unknown proportions, a prior distribution had to be provided for the true proportions. All samples were analysed as potentially including two variants but, to allow for the possibility that only one was present, the prior distribution used had three components: two discrete components, representing the two possible “pure” cases, plus a continuous component representing the possibility of an actual mixture, using an uninformative Beta distribution. Fitting the model with this non-standard prior distribution for proportions used a combination of custom-written code in R and JAGS, and produced a posterior distribution in the same form. The credible intervals used to summarise the posterior distributions combine these discrete and continuous components, as do the Root Mean Squared Errors calculated when the true proportions are known.

## Results

### Comparison of variant frequency estimation using each sequencing method in synthetic mixes

We tested each sequencing protocol in both the same synthetic mixes at high and low concentrations and in the same set of wastewater samples. Coverage plots can be found in Supplementary Materials Figures 1 and 2 for 1 in 10 and concentrated solutions, respectively.

**Figure 1:**
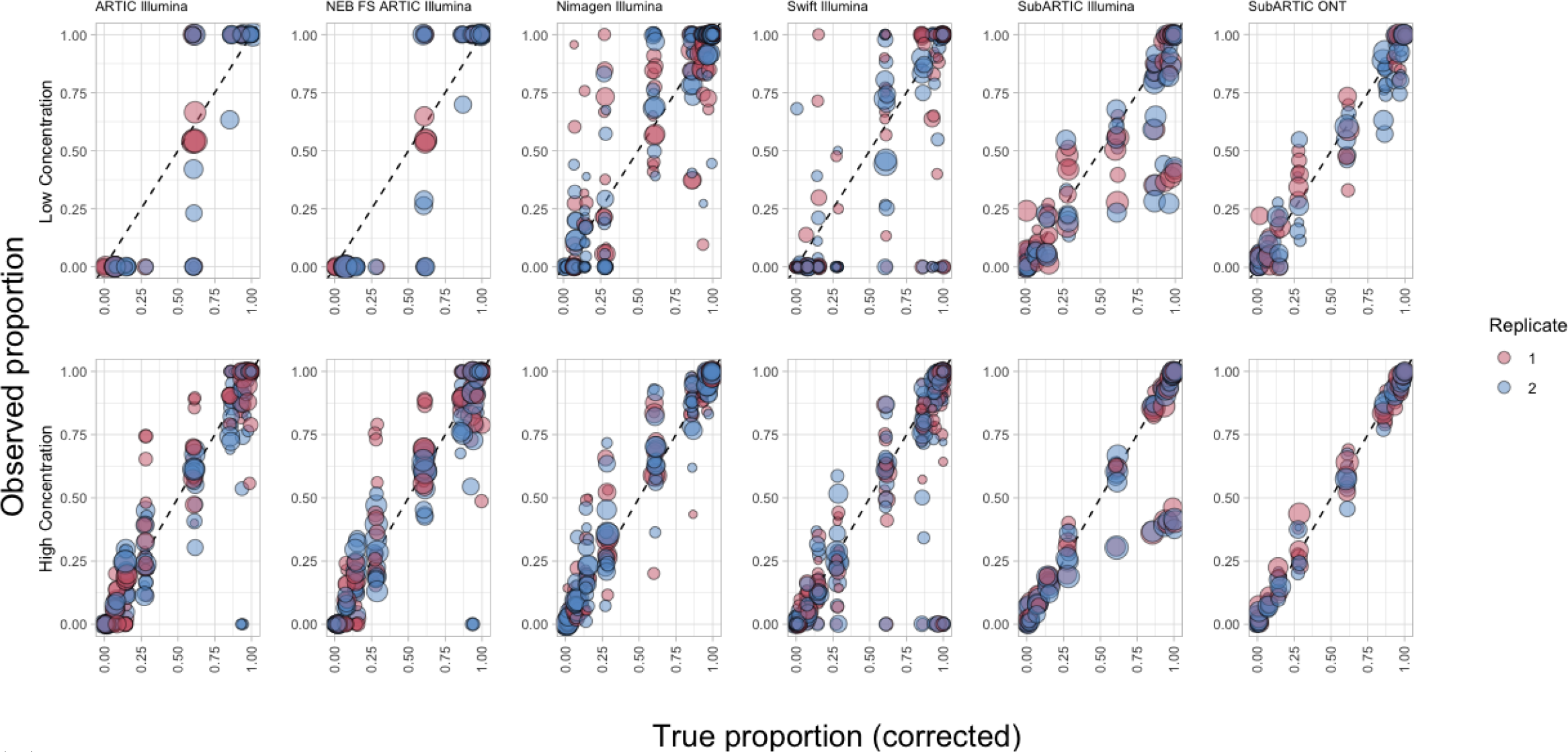
Estimated frequency of each SNP diagnostic of the SHEF variant relative to its expected actual proportion (the 11 corrected frequencies in Table 1) at two concentrations for each of a range of alternative sequencing methods. Two replicates were performed for each method and concentration. Points are weighted by size based on the total number of reads. Points are more opaque where they overlie each other. Sequence reads were not always obtained for every SNP at every frequency.

**Figure 2:**
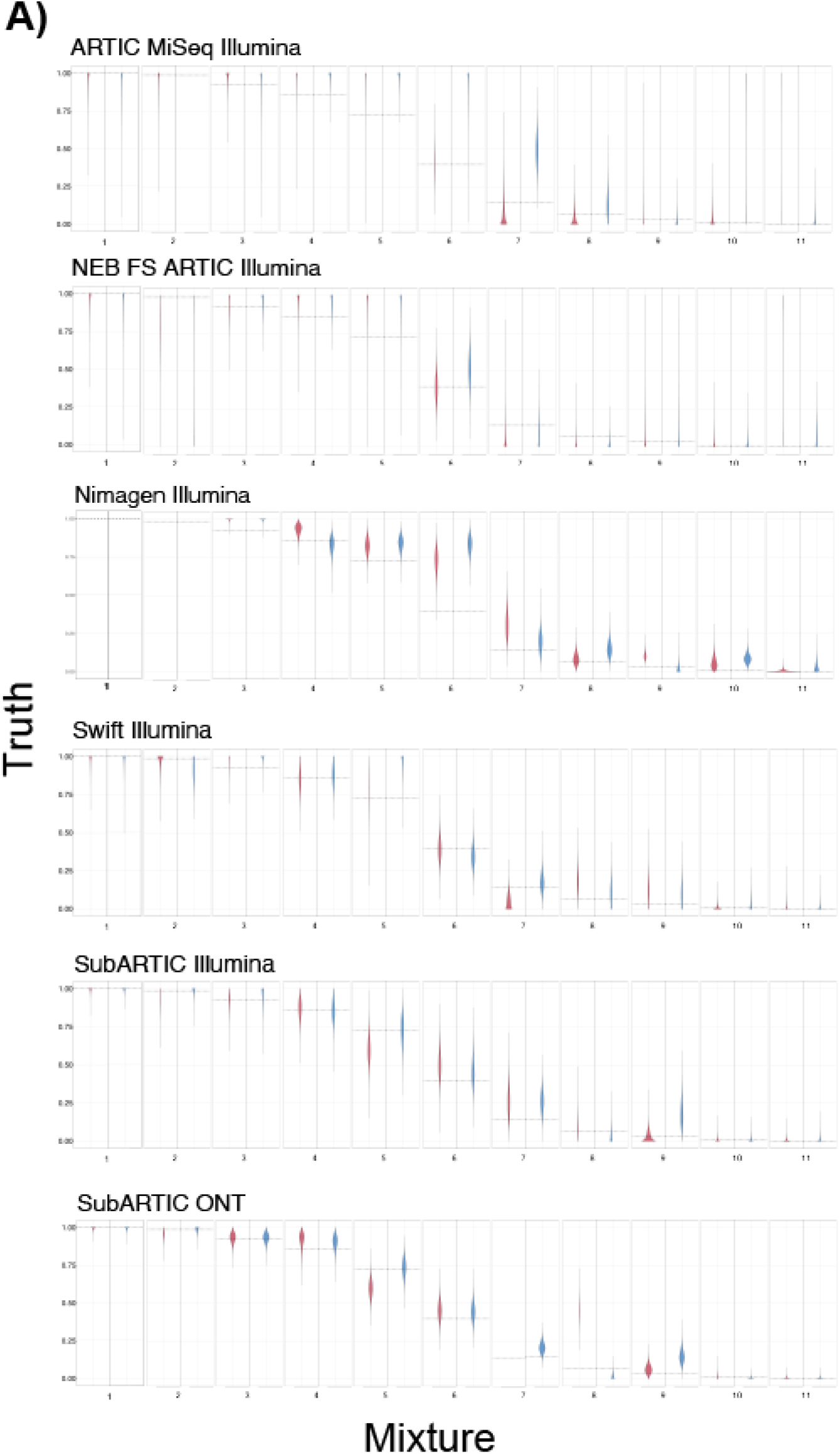
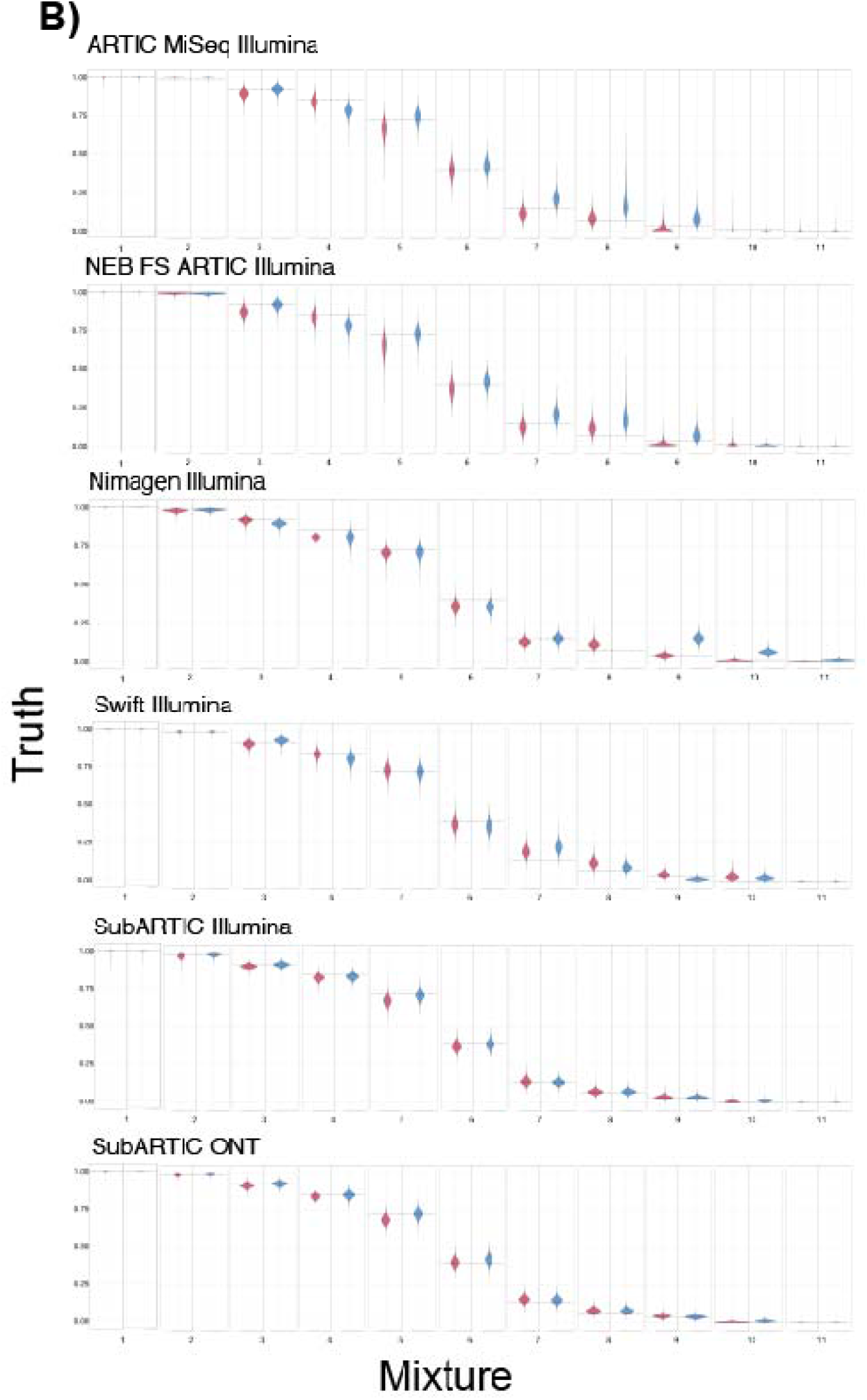
Estimation of variant frequency (truth) shown relative to expectation in (a) 1/10 dilution and (b) concentrated synthetic mixes, using alternative sequencing methods. For each method we show the overall estimate and 95% credible intervals. The known proportion of variant frequency at each mixture is indicated by a dashed horizontal line.

We sequenced each synthetic RNA mixture in duplicate and compared the frequency estimate of each SNP between replicates (Supplementary Materials Figure 3a and 3b). In general, the frequency estimates were highly consistent between replicate runs within a sequencing method (Figure 1 and Figure 4).

**Figure 3:**
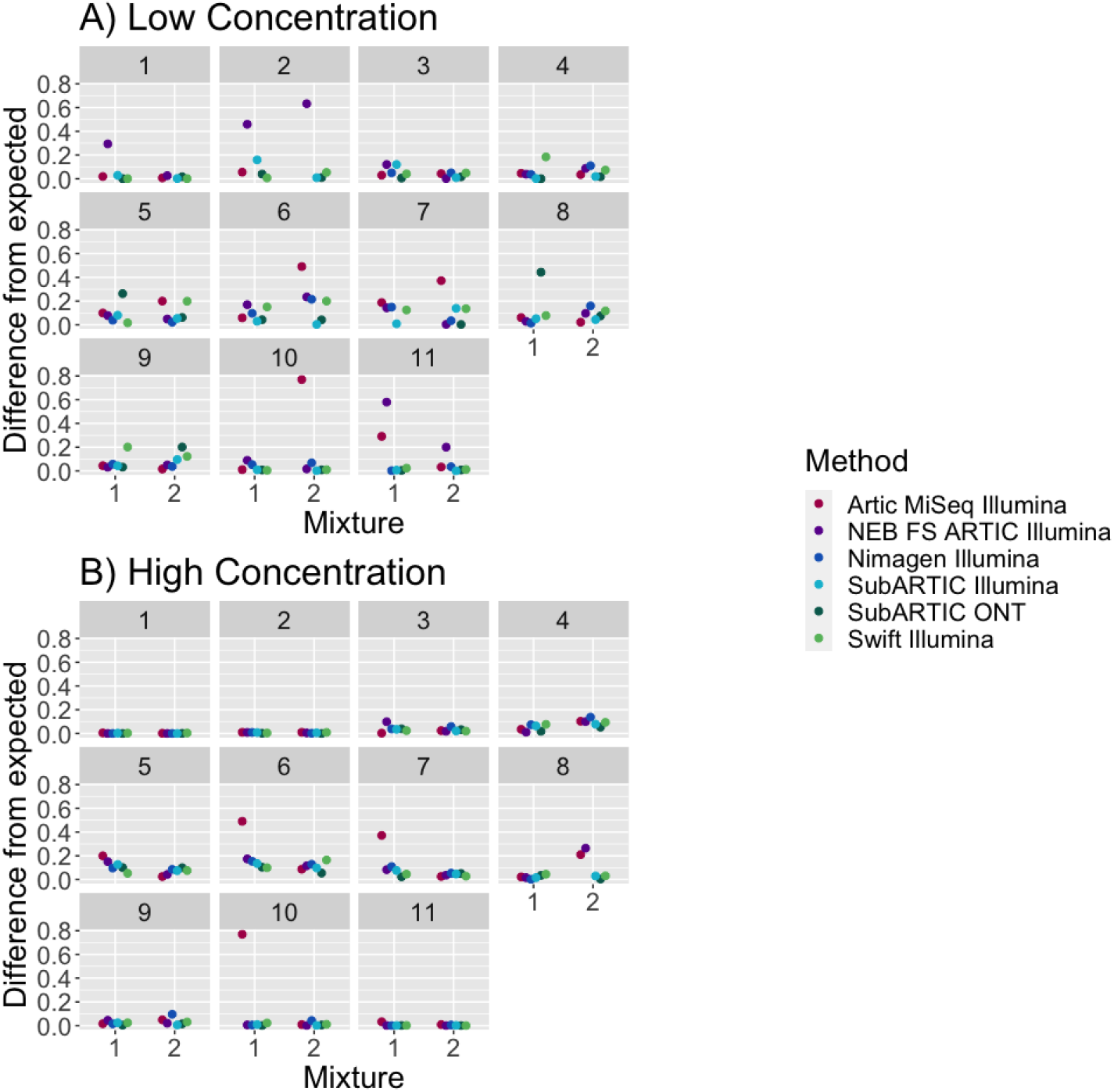
The absolute difference between the predicted and expected variant frequency for the synthetic mixture when either (a) in a 1/10 dilution or (b) concentrated. The predicted variant frequency was calculated as the posterior mode. Missing points are the result of models failing to run due to low read counts.

**Figure 4:**
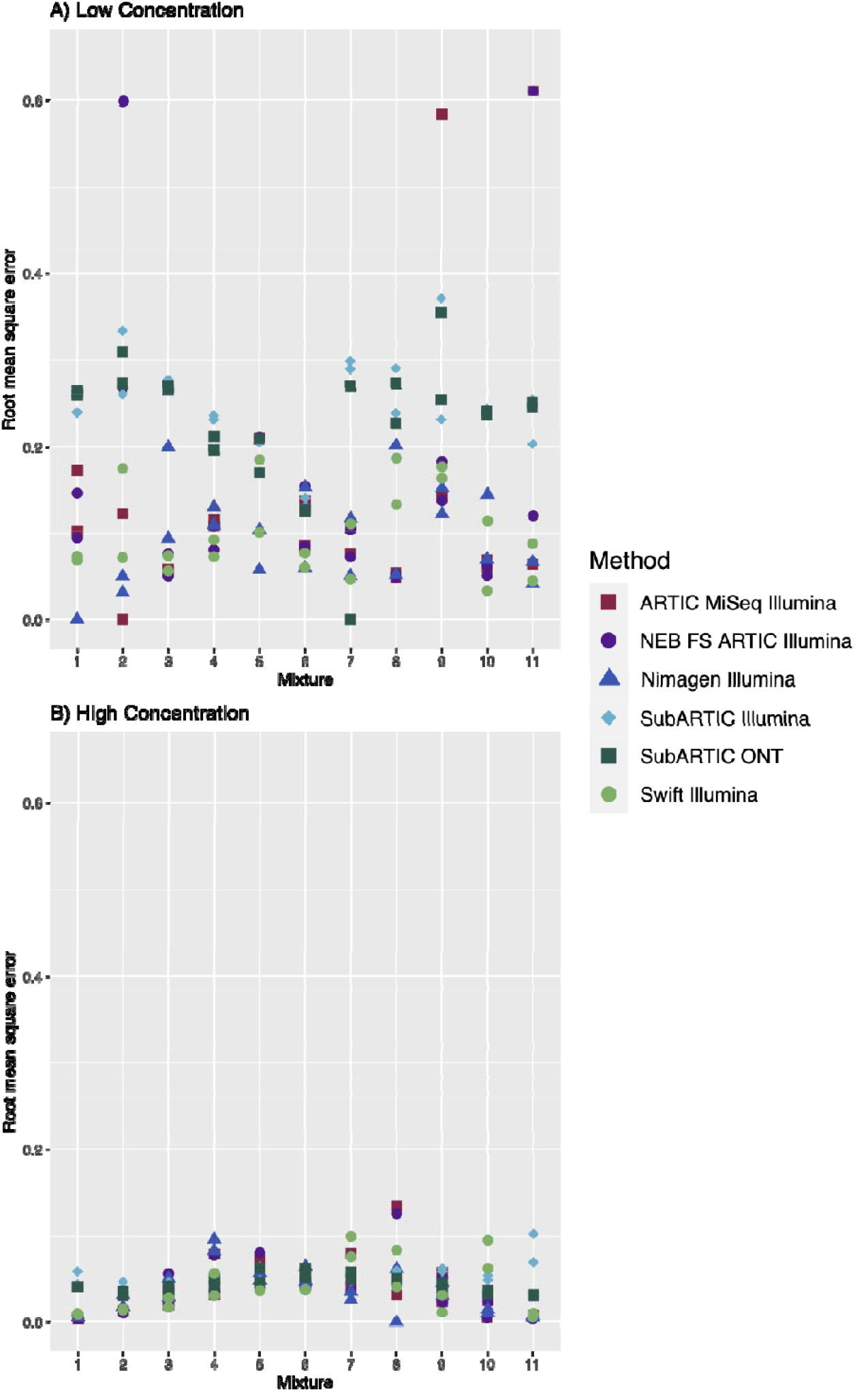
The root mean square error for each method for the synthetic mixture when either (a) in a 1/10 dilution or (b) concentrated. Duplicate points represent the two replicates. A table of raw values can be found in the Supplementary Materials Table 3 and 4.

Spurious (non-variant) SNPs were detected consistently between replicates at usually low frequency (<10%) in each experiment. Such SNPs are unlikely to affect variant frequency estimates, given that the proportion of affected SNPs is very small and each variant is characterised by multiple SNPs (6 SNPs in the spike region and 13 SNPs for whole genome were used here, Table 3). By contrast, such SNPs detected using Illumina and ONT tended not to be consistent with each other (Supplementary Figure 3), illustrating that, while spurious SNPs were nonrandom within a method, they tended to differ between methods.

**Table 3:** Nucleotide changes used in identification of variants. SubARTIC protocols used the spike only region, whereas Illumina protocols were whole genome.

| Spike protein only nucleotide | Whole genome nucleotide |
| --- | --- |
| A23063T | A23063T |
| C23271A | C23271A |
| C23604A | C23604A |
| C23709T | C23709T |
| C23731T | C23731T |
| T24506G | T24506G |
|  | C3267T |
|  | C5388A |
|  | T6954C |
|  | C27972T |
|  | G28048T |
|  | A28111G |
|  | GAT28280CTA |

**Table 4:** Method performance values calculated as the (i) average (mean) absolute difference between estimated variant frequency (truth) and known concentration of variants (difference from expected) and (ii) average (mean) root mean square error across all mixtures and replicates for each method.

| Method | Concentration | Difference from expected | Root mean square error |
| --- | --- | --- | --- |
| ARTIC Illumina | Low | 0.138 | 0.145 |
| ARTIC Illumina | High | 0.114 | 0.378 |
| NEB FS ARTIC Illumina | Low | 0.156 | 0.161 |
| NEB FS ARTIC Illumina | High | 0.055 | **0.038** |
| Nimagen Illumina | Low | 0.068 | 0.092 |
| Nimagen Illumina | High | 0.053 | **0.036** |
| Swift Illumina | Low | 0.082 | 0.101 |
| Swift Illumina | High | 0.040 | **0.039** |
| SubARTIC Illumina | Low | 0.042 | 0.246 |
| SubARTIC Illumina | High | 0.039 | 0.056 |
| SubARTIC ONT | Low | 0.062 | 0.229 |
| SubARTIC ONT | High | 0.030 | **0.044** |

### Estimation of variant frequencies

The frequencies of each SNP diagnostic for each synthetic RNA variant are shown for each mixture and method at both concentrations (Figure 1). The noise in the estimates clearly increases at lower concentration in each case, with appreciable dropout of amplification and sequencing for several sequencing protocols at low concentrations, especially at low frequencies.

The SubARTIC ONT method appears to show the tightest apparent correlation with expectation at high concentration, but is based on fewer SNPs than the whole-genome assays (Figure 1). We therefore used statistical modelling to formally compare overall variant frequency estimations across the sequencing datasets (Figure 2).

As expected, the high concentration mixtures performed better than the low concentration mixtures in terms of both how well variant frequency was predicted (Figure 2 and 3, and Table 4) and the amount of variance in the posterior distribution of predicted variant frequency (Figure 2 and 4, and Table 4).

SubARTIC ONT indeed performed best (determined by performance in average difference from expected and RMSE) in high concentration mixtures (Table 4) and Nimagen Illumina performed best in low concentration mixtures (Table 4). At high concentrations, SubARTIC ONT, Nimagen Illumina, Swift Illumina and SubARTIC Illumina performed comparably well (Table 4). ARTIC Illumina performed less well at high concentrations. At low concentrations, Nimagen Illumina was the best at predicting variant frequencies and had the lowest root mean square error (Table 4). SubARTIC Illumina, Swift Illumina, SubARTIC ONT, ARTIC MiSeq Illumina and NEB FS ARTIC Illumina were able to give reasonable estimates of variant frequencies but had large variance in the prior distribution.

### Wastewater sampling

All methods were comparable in predicting variant frequency in wastewater samples (Figure 5). All methods show the samples were dominated by the main variant (Alpha) in circulation at the time in line with clinical data.

**Figure 5:**
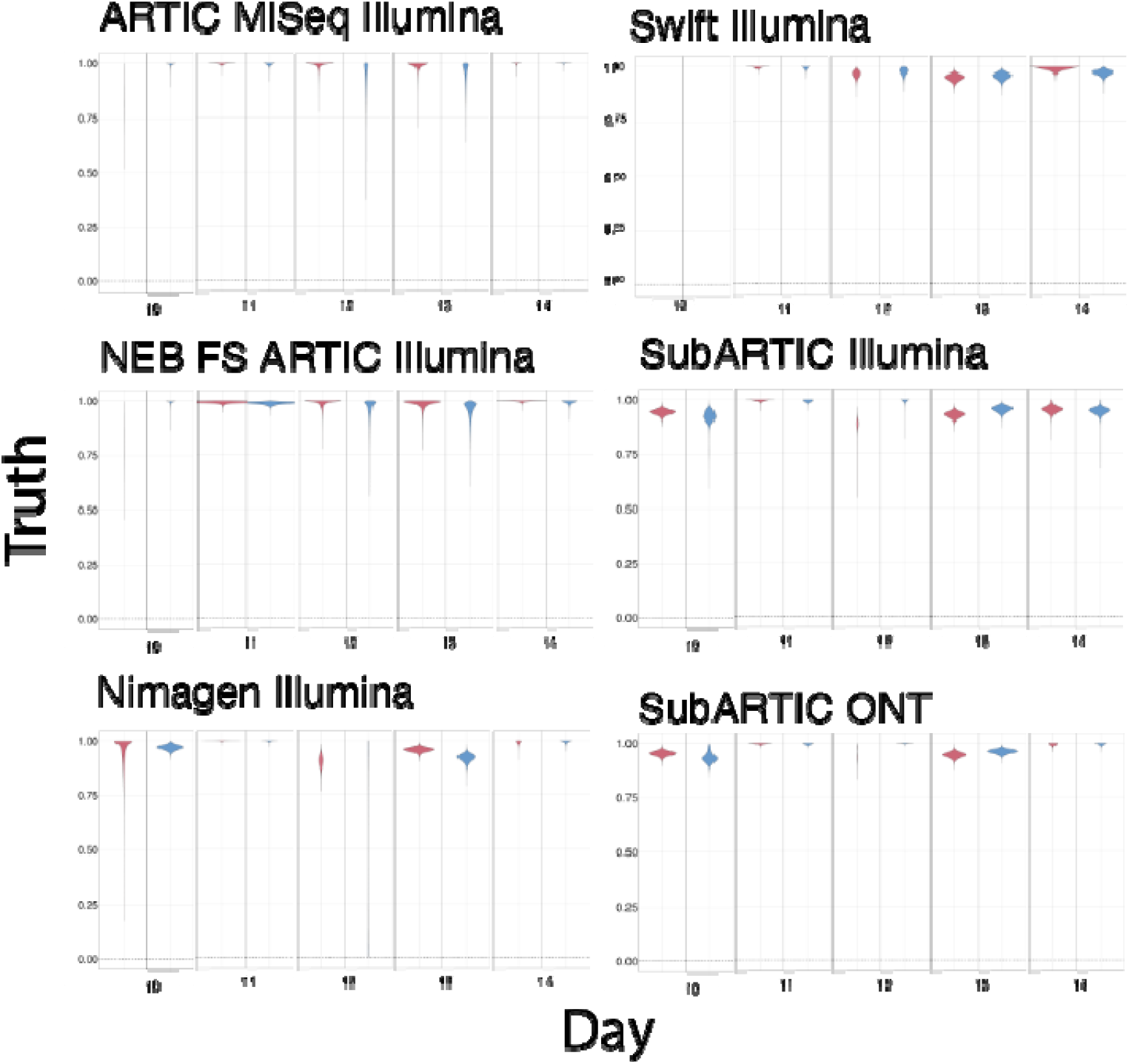
Estimation of variant frequency (truth) in wastewater samples over 5 days in London, UK, in January 2021 using alternative sequencing methods. For each method we show the overall estimate and 95% credible intervals. The different colours represent the two replicates used in this analysis.

## Discussion

Our study has shown that different sequencing methods can detect variant frequency. Though some methods performed better than others, when at high concentrations all methods showed a similar degree of accuracy in determining variant levels. This is especially true when mixtures were dominated by one variant, as indicated in mixtures 1, 2, 10 and 11 (see Figure 3). For these mixtures, there is also less variability around predicting variant frequencies (Fig. 4). ARTIC Illumina, however, had the greatest difference between observed and expected variant frequencies. The lower performance of ARTIC is perhaps unsurprising given the longer insert length: as RNA degrades quickly, the longer insert lengths needed for sequencing mean fewer RNA fragments can be sequenced. However, in the wastewater samples, all the methods were consistent in showing that the frequency of alpha was in the range 90–100%, including ARTIC. However, there is a suggestion that the methods that were more accurate for synthetic mixes are again more precise: all except the ARTIC methods, for example, predict that alpha was actually at 90% on 13th January.

At high concentrations, all other methods (other than ARTIC) performed comparatively similarly, meaning that the methods could be used interchangeably and would not lead to significant differences in estimated variant predictions. ARTIC and NEB FS ARTIC performed less well than the other methods. The SubARTIC protocols had the largest error at low concentrations. As SubARTIC uses sequencing of the Spike region only, the greater coverage (and therefore the additional SNP targets of interest) gained from whole-genome sequencing likely enables better identification of variants when SARS-CoV-2 is at low concentration in a sample. The performance of the two sequencing platforms with the SubARTIC method was comparable at both concentrations. This is somewhat surprising, given that the sequencing error rate in ONT technologies is generally higher than that of Illumina (Delahaye and Nicholas 2021).

It is notable that the most successful methods were those that used short amplicons (∼100–300 bp), despite the input synthetic RNA targets being ca 6,000 bp. We did not test the size of the cDNA produced from these templates but it seems likely that it was similarly large, and far larger than the amplicons used by any of the methods. The results may therefore indicate that, under the conditions of this trial, with many multiplexed targets at low concentration using short amplicons, small amplicons provide a significant benefit simply due to the higher efficiency of amplifying short sequences by PCR.

Finally, the comparison among methods described here used mixtures of synthetic RNA. These RNAs were pure, without the multiple potential contaminants (such as surfactants) that are difficult to remove from RNA extracted from wastewater. Sequencing methods might vary in their sensitivity to such contamination. We attempted to test this by using all the methods to sequence the same set of RNAs extracted from wastewater. All methods were successful at identifying the dominant variant, alpha, but as this was always at very high frequency (>90%), this was not a sensitive test of their respective sensitivity and accuracy for wastewater samples. Such an analysis would require the use of wastewater containing more intermediate frequencies of two or more variants, ideally where the concentration of each was known. This test is currently in progress.

Two of the methods identified and assessed in this study to be successful were subsequently adopted for intensive national wastewater screening programmes: Nimagen for England and Wales, and SubARTIC for Scotland. In conclusion, this study revealed that new-generation sequencing methods, including those that focus only on the Spike region and on both Illumina and Oxford Nanopore platforms, can predict variant frequencies in mixed samples of SARS-CoV-2 with a high degree of accuracy.

## Data Availability

All data produced in the present study are available upon reasonable request to the authors

## Acknowledgements

We thank the Natural Environment Research Council (NERC) for supporting this project (N-WESP, NE/V010441/1 grant to TB) and the NERC Environmental Omics Facility (NEOF).

## CRediT authorship contribution statement

### University of Sheffield

**Paul Blackwell:** Conceptualization, Formal analysis, Investigation, Methodology, Supervision, Writing & editing. **Terry Burke:** Conceptualization, Formal analysis, Funding acquisition, Methodology, Supervision, Writing & editing. **Helen Hipperson:** Methodology, Investigation, Formal analysis. **Gavin Horsburgh**: Methodology, Investigation. **Kathryn Maher:** Conceptualization, Formal analysis, Investigation, Methodology. **Paul Parsons:** Conceptualization, Investigation, Methodology, Writing & editing. *Lucy Winder:* Formal analysis, Writing & editing.

### University of Liverpool

**Claudia Wierzbicki:** Methodology, Investigation, Writing & editing. **Steve Paterson**:

Conceptualization, Supervision, Funding acquisition, Writing & editing.

### University of Exeter

**Aaron Jeffreys:** Methodology, Investigation…

### UKHSA

**Mathew Brown:** Conceptualization, Methodology, Supervision, Writing & editing. **Irene Bassano**: Formal analysis. **Hubert Denise:** Formal analysis, Writing & editing. **Mohammad Khalifa**: Formal analysis. **Aine Fairbrother-Browne:** Formal analysis.

### University of Bangor

**Kata Farcas:** Conceptualization, Methodology, Supervision… **Rachel Williams**: Investigation….

