## Supplementary files for "Comparison of multiple whole-genome and *Spike*-only sequencing protocols for estimating variant frequencies via wastewater-based epidemiology"

**Supplementary Figure 1:** Log mean coverage for each sequencing method at a 1 in 10 dilution mixture. Note SubARTIC Illumina and SubARTIC ONT are *Spike* regions only.

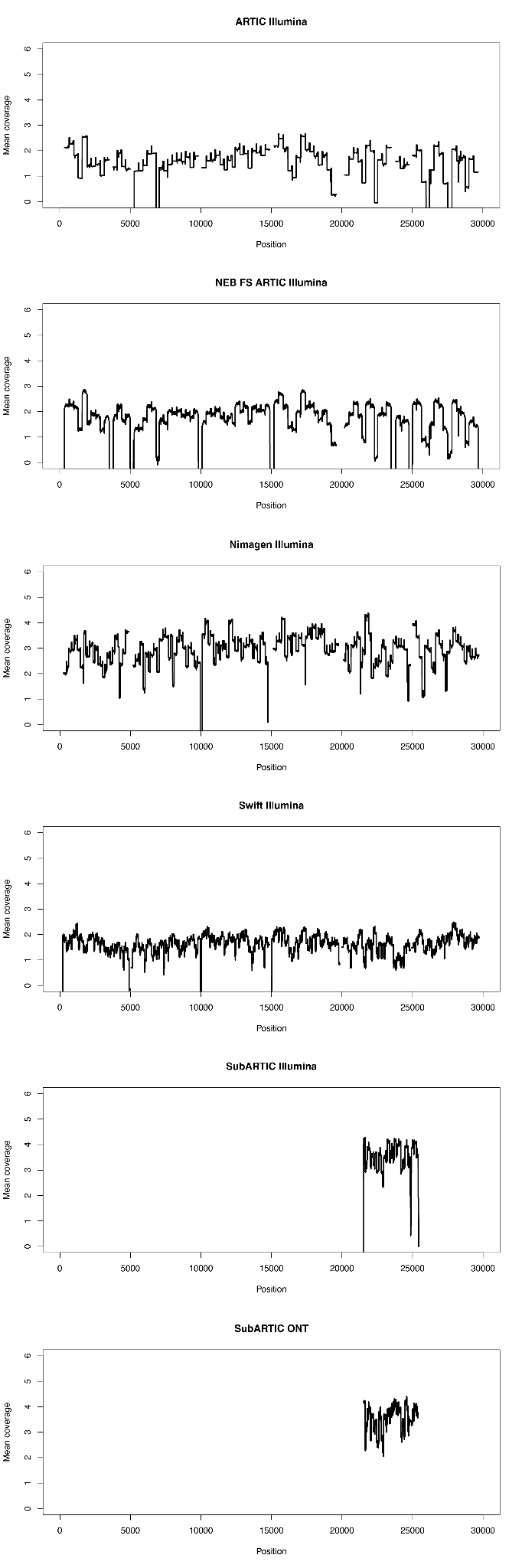

**Supplementary Figure 2:** Log mean coverage for each sequencing method for concentrated solutions. Note SubARTIC Illumina and SubARTIC ONT are *Spike* regions only.

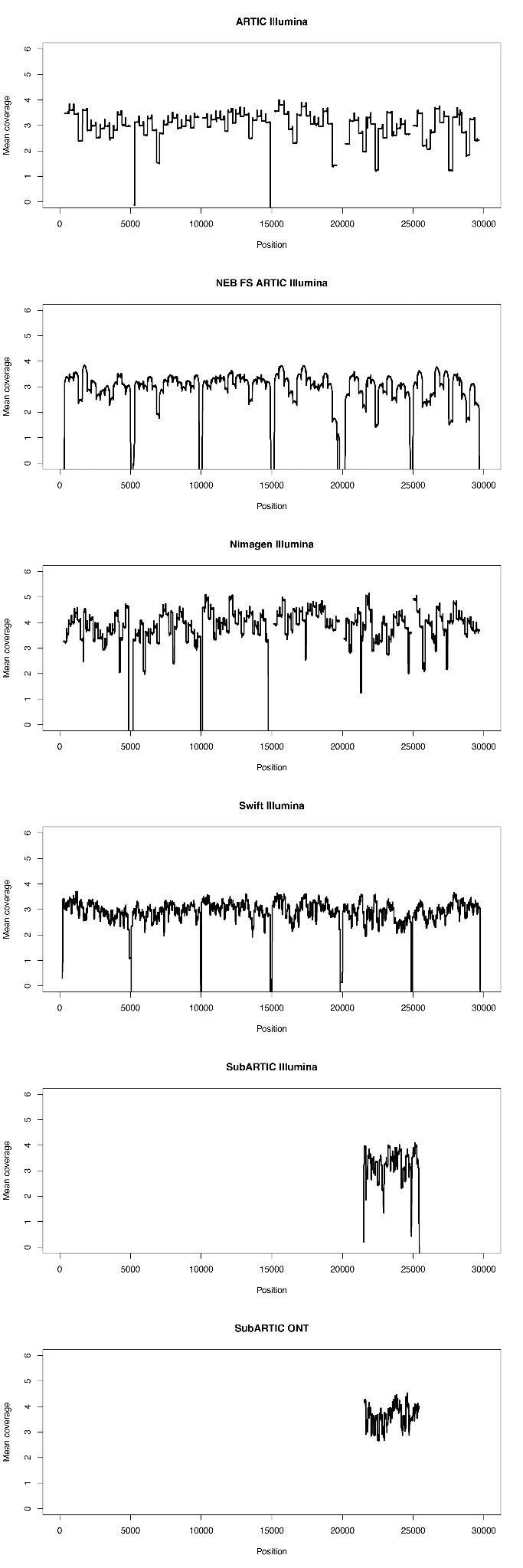

**Supplementary Figure 3:** Frequencies of variant sites in the *Spike* gene for replicate synthetic samples at 0%, 28%, 86% and 100% frequency of the SHEF variant in the mixed sample using the SubARTIC protocol: (A) SNPs and indels detected using Illumina, (B) SNPs (excluding indels) detected using Oxford Nanopore. Replicate samples (R1, R2) are plotted against each other.

A)

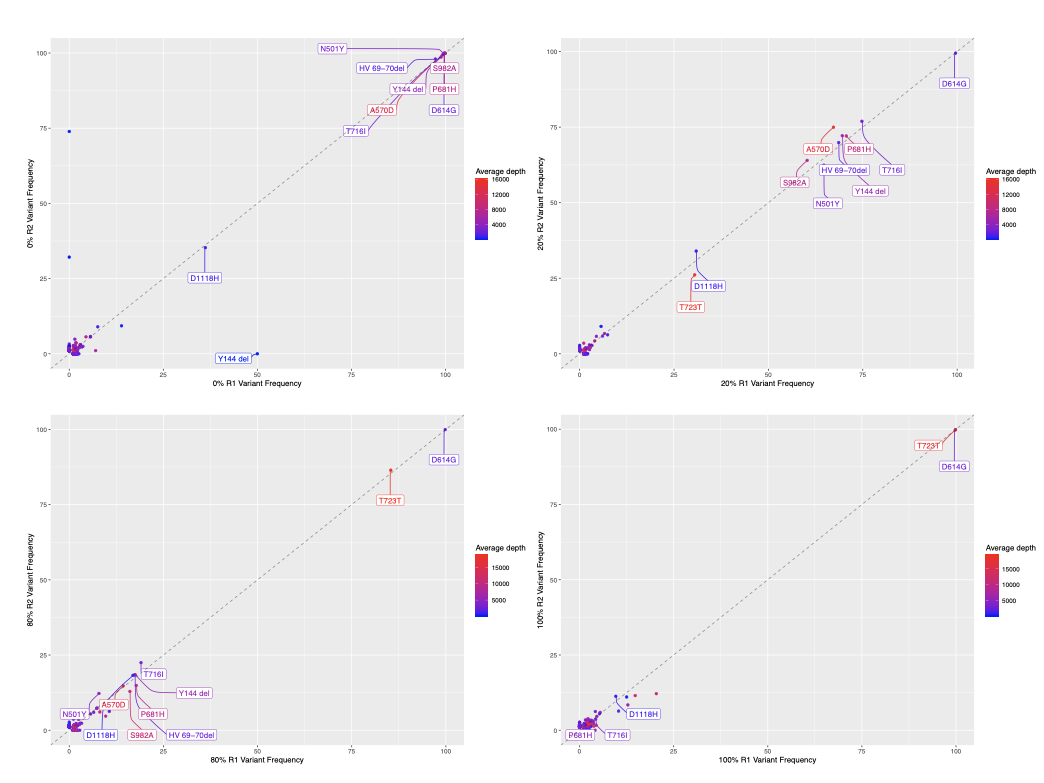

B)

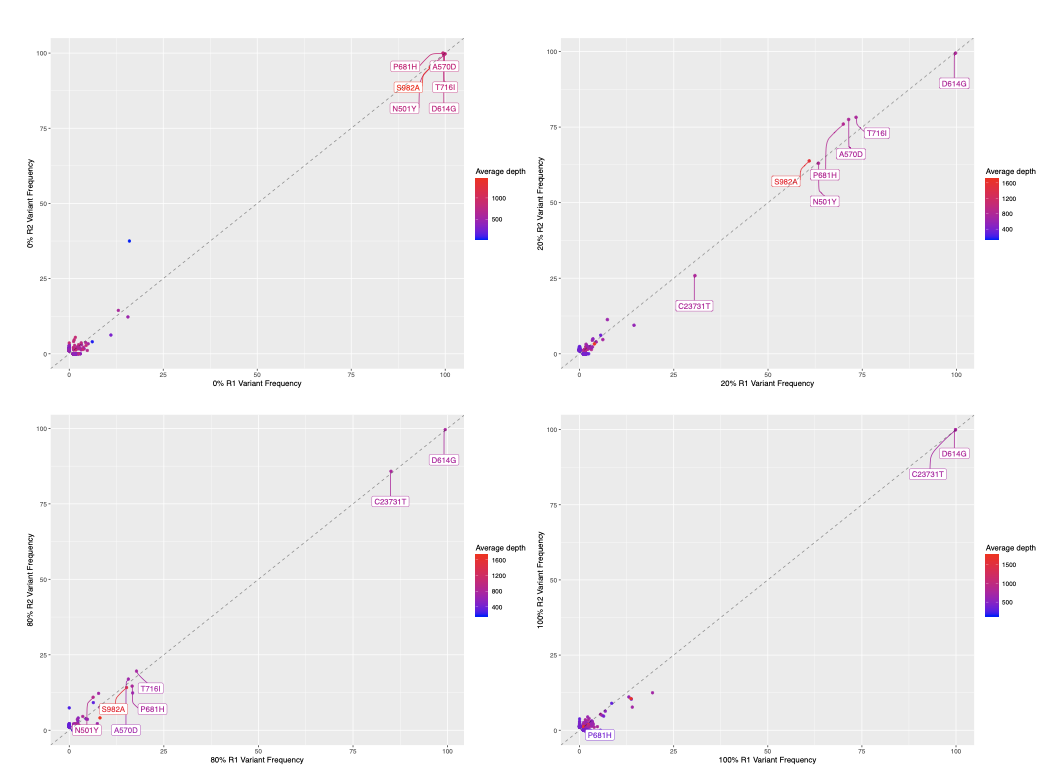

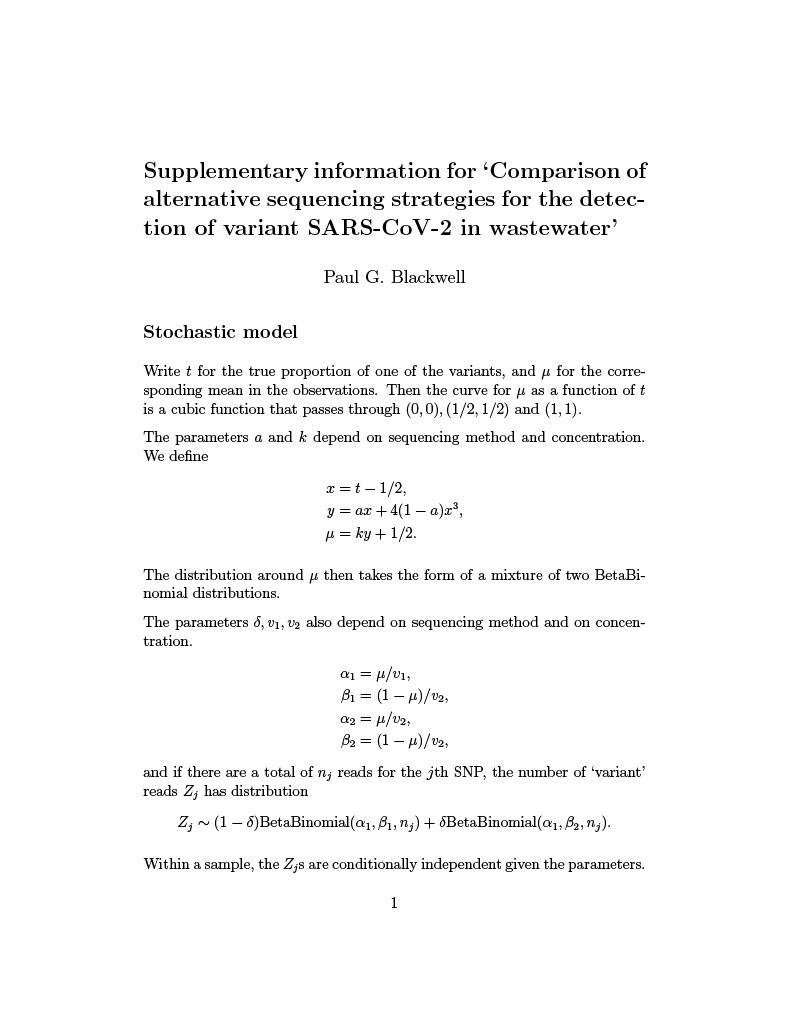

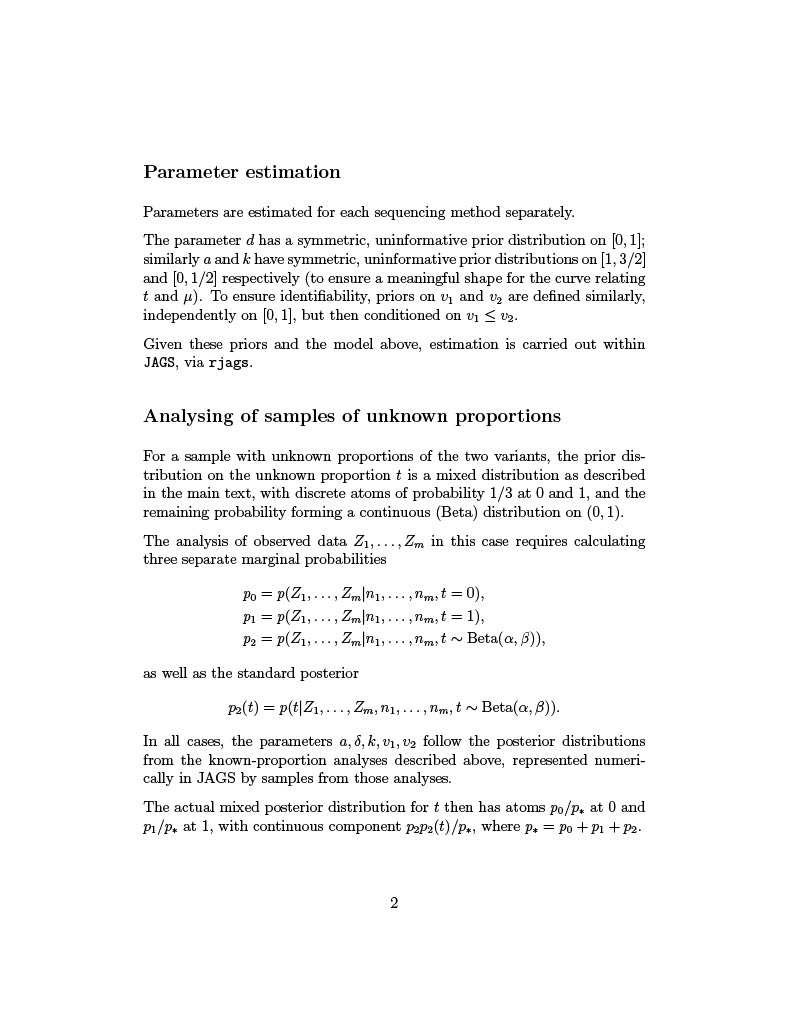

**Supplementary Table 3.** Root mean square error of the results of the MCMC chains for each mixture and method for 1/10 dilution. The results of the MCMC chains are compared to the expected actual variant proportion in the mixture.

|  | Mixture | | | | | | | | | | | | | | | | | | | | | |
| --- | --- | --- | --- | --- | --- | --- | --- | --- | --- | --- | --- | --- | --- | --- | --- | --- | --- | --- | --- | --- | --- | --- |
|  | 1 | | 2 | | 3 | | 4 | | 5 | | 6 | | 7 | | 8 | | 9 | | 10 | | 11 | |
|  | Replicate | | | | | | | | | | | | | | | | | | | | | |
| Method | 1 | 2 | 1 | 2 | 1 | 2 | 1 | 2 | 1 | 2 | 1 | 2 | 1 | 2 | 1 | 2 | 1 | 2 | 1 | 2 | 1 | 2 |
| ARTIC Illumina | 0.102 | 0.171 | 0.120 | - | 0.058 | 0.134 | 0.109 | 0.116 | 0.209 | 0.242 | 0.085 | 0.510 | 0.108 | 0.361 | 0.055 | 0.102 | 0.144 | 0.037 | 0.068 | 0.607 | 0.615 | 0.065 |
| NEB FS ARTIC Illumina | 0.095 | 0.148 | 0.254 | 0.600 | 0.071 | 0.053 | 0.108 | 0.097 | 0.209 | 0.209 | 0.089 | 0.153 | 0.107 | 0.086 | 0.054 | 0.051 | 0.139 | 0.188 | 0.062 | 0.049 | 0.613 | 0.101 |
| Nimagen v2 Illumina | - | - | - | - | 0.069 | 0.069 | 0.083 | 0.054 | 0.111 | 0.124 | 0.338 | 0.438 | 0.190 | 0.081 | 0.039 | 0.093 | 0.076 | 0.026 | 0.076 | 0.085 | 0.010 | 0.043 |
| Nimagen v3 Illumina | 0.613 | 0.614 | 0.602 | 0.635 | 0.553 | 0.648 | 0.532 | 0.531 | 0.420 | 0.419 | 0.350 | 0.370 | 0.500 | 0.502 | 0.553 | 0.490 | 0.589 | 0.587 | 0.608 | 0.552 | 0.577 | 0.614 |
| Swift Illumina | 0.055 | 0.061 | 0.057 | 0.106 | 0.052 | 0.061 | 0.074 | 0.068 | 0.124 | 0.229 | 0.071 | 0.081 | 0.097 | 0.065 | 0.135 | 0.080 | 0.117 | 0.096 | 0.021 | 0.050 | 0.043 | 0.032 |
| SubARTIC Illumina | 0.019 | 0.018 | 0.085 | 0.024 | 0.052 | 0.054 | 0.064 | 0.066 | 0.150 | 0.084 | 0.139 | 0.105 | 0.153 | 0.136 | 0.065 | 0.046 | 0.032 | 0.169 | 0.018 | 0.019 | 0.018 | 0.026 |
| SubARTIC ONT | 0.012 | 0.014 | 0.044 | 0.017 | 0.033 | 0.031 | 0.079 | 0.064 | 0.139 | 0.056 | 0.079 | 0.072 | - | 0.072 | 0.385 | 0.052 | 0.042 | 0.120 | 0.020 | 0.007 | 0.009 | 0.009 |

**Supplementary Table 4.** Root mean square error of the results of the MCMC chains for each mixture and method for the concentrated dilutions. The results of the MCMC chains are compared to the expected actual variant proportion in the mixture.

|  | Mixture | | | | | | | | | | | | | | | | | | | | | |
| --- | --- | --- | --- | --- | --- | --- | --- | --- | --- | --- | --- | --- | --- | --- | --- | --- | --- | --- | --- | --- | --- | --- |
|  | 1 | | 2 | | 3 | | 4 | | 5 | | 6 | | 7 | | 8 | | 9 | | 10 | | 11 | |
|  | Replicate | | | | | | | | | | | | | | | | | | | | | |
| Method | 1 | 2 | 1 | 2 | 1 | 2 | 1 | 2 | 1 | 2 | 1 | 2 | 1 | 2 | 1 | 2 | 1 | 2 | 1 | 2 | 1 | 2 |
| ARTIC Illumina | 0.006 | 0.003 | 0.014 | 0.014 | 0.039 | 0.019 | 0.033 | 0.079 | 0.079 | 0.041 | 0.043 | 0.045 | 0.041 | 0.077 | 0.032 | 0.139 | 0.025 | 0.058 | 0.025 | 0.006 | 0.007 | 0.008 |
| NEB FS ARTIC Illumina | 0.003 | 0.002 | 0.010 | 0.008 | 0.060 | 0.022 | 0.038 | 0.080 | 0.093 | 0.037 | 0.051 | 0.042 | 0.039 | 0.073 | 0.058 | 0.127 | 0.023 | 0.049 | 0.018 | 0.004 | 0.004 | 0.004 |
| Nimagen v2 Illumina | 0.001 | 0.001 | 0.012 | 0.008 | 0.018 | 0.034 | 0.056 | 0.061 | 0.032 | 0.036 | 0.048 | 0.050 | 0.026 | 0.021 | 0.044 | - | 0.011 | 0.113 | 0.005 | 0.052 | 0.001 | 0.010 |
| Nimagen v3 Illumina | 0.004 | 0.005 | 0.903 | 0.012 | 0.095 | 0.136 | 0.074 | 0.100 | 0.085 | 0.534 | 0.083 | 0.066 | 0.047 | 0.147 | 0.027 | 0.558 | 0.585 | 0.584 | 0.608 | 0.613 | 0.005 | 0.611 |
| Swift Illumina | 0.002 | 0.003 | 0.010 | 0.010 | 0.022 | 0.018 | 0.027 | 0.052 | 0.033 | 0.031 | 0.038 | 0.045 | 0.061 | 0.092 | 0.059 | 0.031 | 0.019 | 0.018 | 0.032 | 0.022 | 0.002 | 0.003 |
| SubARTIC Illumina | 0.010 | 0.006 | 0.014 | 0.008 | 0.020 | 0.013 | 0.030 | 0.025 | 0.050 | 0.026 | 0.028 | 0.024 | 0.020 | 0.016 | 0.016 | 0.016 | 0.014 | 0.012 | 0.007 | 0.009 | 0.002 | 0.003 |
| SubARTIC ONT | 0.003 | 0.002 | 0.007 | 0.010 | 0.019 | 0.013 | 0.025 | 0.019 | 0.043 | 0.024 | 0.027 | 0.039 | 0.026 | 0.023 | 0.022 | 0.022 | 0.017 | 0.012 | 0.005 | 0.007 | 0.001 | 0.002 |
